# Spatial modeling of risk factors for human exposure to West Nile virus in rural Madagascar

**DOI:** 10.64898/2026.10.19.26364157

**Authors:** Mauricianot Randriamihaja, Lova Tsikiniaina Rasoloharimanana, Sruthi Veeragandham, Ann C. Miller, Karen E. Finnegan, Sandra Bos, Benedicte Razafinjato, Ezra Mitsinjoniala, Jacques Aubin Kotchofa, Pierre Garneret, Hobisoa L. Razanadranaivo, Felana Angella Ihantamalala, Rado J.L. Rakotonanahary, Veronique Chevalier, Matthew H. Bonds, Matthieu Schoenhals, Michelle V. Evans, Stephen J. Popper, Andres Garchitorena

**Author notes:** These authors contributed equally to this work (Stephen J. Popper, Andres Garchitorena).

## Abstract

**Background:** West Nile virus (WNV) is an emerging disease transmitted from bird hosts to humans by Culex mosquitoes. Thousands of human cases of WNV infection are reported each year in the United States and Europe, and circulation has been confirmed in Africa and the Indian Ocean, including Madagascar. However, little is known about the current riskscape for people in Madagascar, and what determines its local distribution and human exposure across the population. The goal of this study was to assess WNV seroprevalence and identify associated socio-ecological risk factors in rural Madagascar.

**Methodology and principal findings:** A cross-sectional serological survey was conducted in Ifanadiana District in 2021, nested within a representative longitudinal cohort. We evaluated serological responses to WNV in 3,948 individuals using a multiplex bead assay. We estimated WNV seroprevalence using a Gaussian Mixture Model (GMM) and characterized environmental, socioeconomic, and demographic factors that could be associated with exposure to WNV using various data sources, including optical and Synthetic Aperture Radar (SAR) satellite images, climate variables, and household survey data. Overall seroprevalence was 43%, and populations living in geographically isolated areas and those engaged in agricultural activities were at higher risk, while proximity to dense forests and residential areas were protective against WNV exposure.

**Conclusions:** This study found high rates and local heterogeneity in WNV exposure in a rural district of Madagascar. In addition, environmental, climatic, and socio-demographic data were determinants of WNV transmission, providing a valuable tool for surveillance and decision-making in arbovirus prevention and control. Extending this approach to other orthoflaviviruses could improve understanding of local viral co-circulation and promote the development of more precise and context-specific prevention strategies.

## Introduction

Following the first documented large outbreak in the Northern Hemisphere in 1999, West Nile virus (WNV), a mosquito-borne *orthoflavivirus*, became established in the United States and Europe, where thousands of cases are reported each year (Ronca et al., 2021; Abbas et al., 2025; European Centre for Disease Prevention and Control (ECDC) & Authority (EFSA), 2025; Gould et al., 2025). While most infections are asymptomatic, about 20% of WNV infections result in flu-like symptoms such as mild fever, and 1% of infections lead to severe illness in the form of encephalitis, meningitis, acute flaccid paralysis, and, in rare cases, death. Despite attention and surveillance efforts being concentrated in the Northern Hemisphere, WNV circulation is also well documented in Africa and the Indian Ocean region (M. L. Tantely et al., 2016; Cardinale et al., 2017; Mencattelli et al., 2022; Moir et al., 2025). Though ecological conditions in the tropics are especially amenable to arbovirus transmission, lower rates of surveillance and diagnosis leave open questions about its distribution. Some studies have shown seroprevalence rates over 50% among certain populations, indicating WNV may be responsible for an unknown burden of morbidity and mortality (Sule et al., 2018; Broban et al., 2023; Rakotomalala et al., 2023). Understanding the factors associated with the risk of WNV infection in humans is foundational to informing surveillance and control efforts in these regions.

The transmission cycle of WNV is based on a complex network involving domestic and wild birds, which act as reservoirs and amplifiers, and *Culex* mosquitoes, the main vectors responsible for transmission to humans and other dead-end hosts, notably horses (Hayes & Gubler, 2006; Giesen et al., 2023). Given the existence of an animal reservoir and a mosquito vector, the spatio-temporal dynamics of the disease are modulated by a complex interaction of environmental, socio-economic, and demographic factors, which determine the risk of exposure and explain variation observed between areas (Paz, 2015; Hernandez et al., 2019; Giesen et al., 2023). Studies have revealed notable correlations with various environmental factors associated with high WNV infection rates, such as climatic conditions (e.g., temperature and precipitation), landcover (e.g., Normalized Difference Vegetation Index-NDVI, deforestation), soil moisture, and road infrastructure (García-Carrasco et al., 2023; Giesen et al., 2023; Gonçalves do Amaral et al., 2024; Taheri et al., 2025). Some studies have also shown that socioeconomic and demographic parameters significantly influence exposure to WNV at the household level, including occupation, cultivated land, livestock farming, economic status, and education (García-Carrasco et al., 2023; Odigie et al., 2024; Vargas Campos et al., 2025), as well as open defecation, insecticide spraying, and water source (Zinsstag et al., 2018; WHO and UNICEF, 2021; Klingelhöfer et al., 2023; Gonçalves do Amaral et al., 2024). Additionally, exposure to WNV can be influenced by individual-level factors such as the use of mosquito nets, age, and gender (Kalantari et al., 2019; Solanke et al., 2023).

Most existing studies examining the socio-ecological drivers of WNV were conducted at a national or multi-country level (Mencattelli et al., 2022; Giesen et al., 2023), and rarely at the sub-national level, where local socioeconomic and climatic conditions may lead to substantial spatial heterogeneity in WNV risk and exposure. Moreover, research to date has been mostly focused on the Northern Hemisphere, where WNV outbreaks are primarily urban or suburban (Klingelhöfer et al., 2023), leaving substantial knowledge gaps in the spatial distribution in rural areas of the tropics and sub-tropics. National seroprevalence of WNV in Madagascar is estimated to be over 25% (Broban et al., 2023; Rakotomalala et al., 2023), with rural areas experiencing nearly double the seroprevalence of urban areas (Broban et al., 2023), but these studies had very limited data available from any single region or district. Here, we examine local heterogeneity in WNV exposure and the associated risk factors in the rural district of Ifanadiana, in southeastern Madagascar. We integrate socio-economic and serological data from a population-representative household survey conducted in 2021 with environmental variables from satellite imagery, climate models, and other sources to identify socio-ecological factors associated with individual-level exposure to WNV. This approach aims to refine our understanding of local transmission dynamics, identify spatial risk factors, and guide the development of targeted prevention and control strategies against WNV and other orthoflaviviruses in low-resource settings.

## Methods

### Study area

The study was conducted in Ifanadiana, a rural district of approximately 200,000 people located in the Vatovavy region, in southeastern Madagascar. Ifanadiana has been part of a district-wide health system strengthening initiative in partnership between the government of Madagascar and an international NGO (Cordier et al., 2020; Garchitorena et al., 2025). This partnership, focused on expanding access to essential health services and reducing preventable mortality, also provides unique population-level household data. The district covers an area of approximately 3,970 km^2^. On the western side of the district is a protected rainforest park that hosts over a hundred species of wild birds (Madagascar National Parks, 2025). The district has a single paved road running east-west, with tertiary (unpaved) roads connecting the northern and southern axes of the district (Figure 1). The district has a humid tropical climate, with an average annual rainfall of 1,200 mm and an average annual temperature between 15 and 28 °C. Ifanadiana is characterized by a matrix of over 416 km² of dense forest areas (including the national park) and agricultural land used for rice production, on which more than 80% of the population depends. The terrain is mountainous, with an elevational gradient from 1,400m in the west to 100m on its eastern side. In addition, the population experiences high rates of poverty. Only 14.9% of the population has access to an improved source of drinking water, while nearly 50% practice open defecation and 94.8% of households report having at least one mosquito net (Miller et al., 2018). These conditions reflect high health vulnerability and have been shown to be key determinants of transmission of mosquito-borne diseases to humans (Pourtois et al., 2023; Evans et al., 2025).

**Figure 1.**
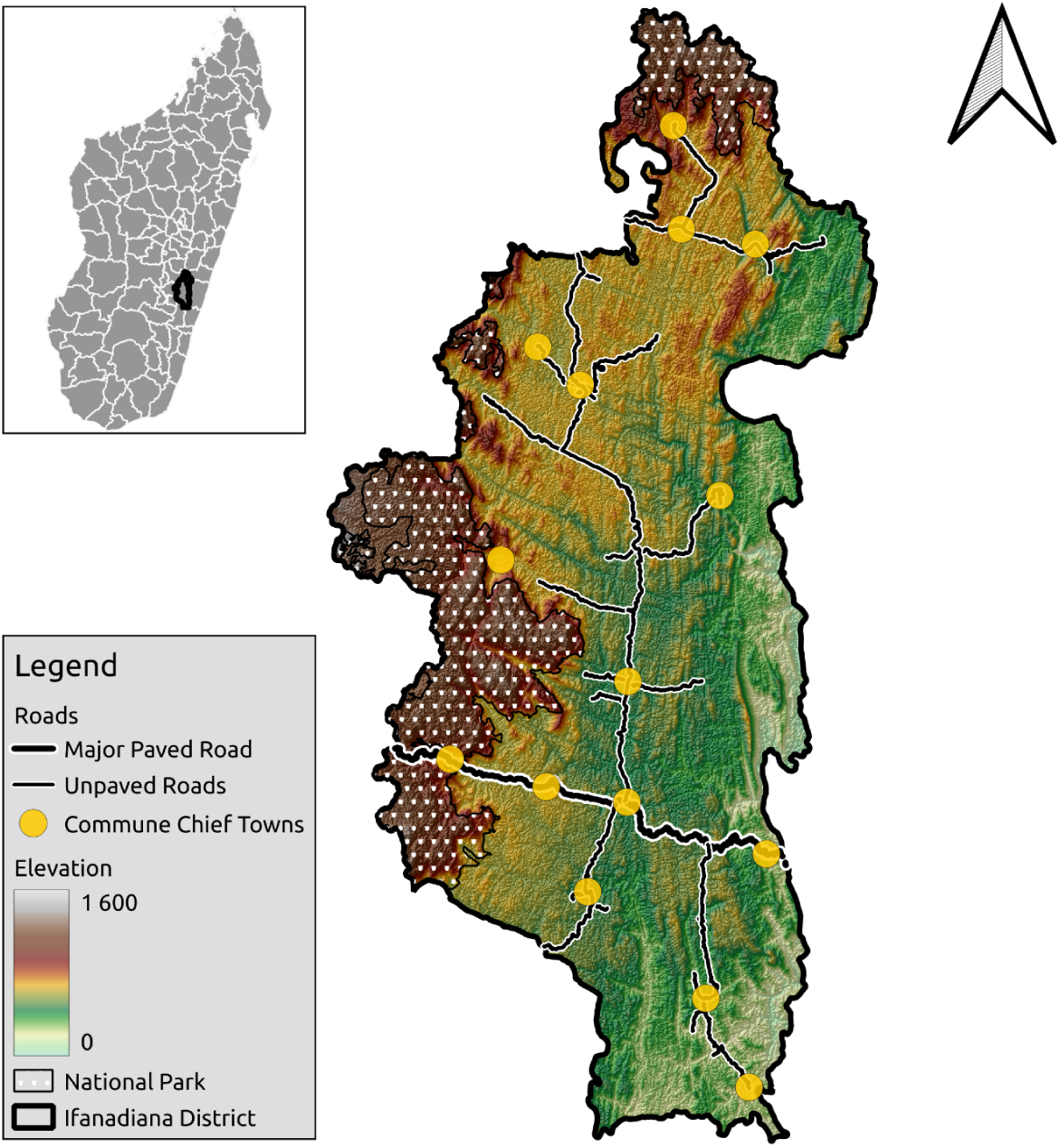
Map of Ifanadiana district, southeastern Madagascar. The inset map shows the location of Ifanadiana District within Madagascar. The main map shows the elevation gradient in the district, along with the limits of Ranomafana National Park (indicated by white symbols), road infrastructure (black), and main towns (yellow). Sources: NASA Earthdata (https://dwtkns.com/srtm30m/) used for elevation data, HumData Madagascar (https://data.humdata.org/dataset/cod-ab-mdg) used for administrative boundaries, Contains information from OpenStreetMap and OpenStreetMap Foundation, which is made available under the Open Database License (https://www.openstreetmap.org/copyright).

## Data collection

### Survey data collection

A population-based survey was conducted from April to June of 2021 as part of the Ifanadiana Health Outcomes and Prosperity longitudinal Evaluation (the “IHOPE” cohort), which allowed us to collect blood samples using dried blood spot (DBS) cards, as well as socioeconomic and demographic data on a sample of households representative of Ifanadiana district. Using a two-stage cluster sampling design, the survey was administered to two strata representing two separate geographic areas defined by whether households were part of the initial group that benefited from the ongoing health system strengthening partnership that was progressively implemented across the district starting in 2014 (Miller et al., 2018). Each stratum consisted of 40 randomly selected clusters, each of which contained 20 randomly selected households, for 1600 households in total. First conducted in 2014, the survey has since been conducted every two years, with the exception of 2020 which was delayed until 2021, with any non-participating households replaced by others from a list generated at the initial randomization. Household and individual questionnaires were adapted from the Demographic and Health Survey (DHS) and administered to eligible household members (men aged 15-59 years and women aged 15-49 years). This included questions regarding socio-demographic, health, and economic information at the individual and household level, as well as anthropometric measurements for all members present at the time of the visit (Miller et al., 2018). The Madagascar National Institute of Statistics (INSTAT), which implements all major national health surveys in the country, was responsible for data collection, survey coordination, training and oversight. In addition, dried blood spots (DBS) were collected by trained nurses on Whatman 903 protein saver cards from a finger-prick capillary blood sample from consenting members of all ages in the surveyed households. A total of 6,636 DBS samples were collected for serological analysis. Five blood spots were obtained for each sample, air-dried, and stored in dry conditions in bags containing desiccants for subsequent analysis.

Serological analyses of dried blood spots (DBS) were performed at the Institut Pasteur de Madagascar using a multiplex bead-based immunoassay (Luminex xMAP technology). DBS were sub-punched into 3-mm disks and eluted overnight at 4°C with agitation in 1mL of PBS-T 0.5% (VWR E404; Sigma P1379). Eluates were then transferred to 1.5 mL tubes and stored at −20°C until analysis. Antigen coupling was performed using the xMAP Antibody Coupling Kit (Luminex, 40-50016). Briefly, 10 µL of Sulfo-NHS and 10 µL of EDC were added to 5×10⁶ washed Magplex™ MC100XX beads followed by the addition of 25 µg of recombinant antigens. All proteins were obtained from the Native Antigen Company (UK). The West Nile virus (WNV) recombinant antigens used were the envelope protein (E; REC31614) and the nonstructural protein 1 (NS1; WNV-NS1). The mixture was incubated for 2 hours in the dark at room temperature with gentle shaking. For the immunoassay 1000 beads per antigen were transferred into each well of a 96-well round-bottom microplate (Corning 3789, Fisher Scientific). 100 µL of undiluted eluates and controls were added and incubated for 45 minutes with agitation. After two washing steps, 100 µL of goat anti-human IgG Fc R-PE (Thermo Fisher, H10104), diluted 1:400, was added and incubated for an additional 45 minutes with agitation. Beads were then resuspended in 120 µL of buffer and read on the Magpix instrument. The instrument identifies each bead region by its internal fluorescent signature and quantifies antibody binding by measuring the mean fluorescence intensity (MFI) of the reporter signal via LED-based excitation.

### Socioeconomic and demographic factors

From the IHOPE survey, we selected socioeconomic and demographic variables at the household and individual levels relevant to WNV transmission (Table 1). Household-level variables included the education level and economic status of the household head (Vargas Campos et al., 2025), as well as information on agricultural occupations and livestock ownership, which may attract *Culex* mosquitoes to non-human mammalian hosts and influence exposure risk during daily outdoor activities (Odigie et al., 2024). Additional household-level variables also included the characteristics of cropland (Erazo et al., 2024), housing quality (Lockaby et al., 2016; Power et al., 2022), the use of unprotected or uncovered water sources that may serve as mosquito habitats (Gonçalves do Amaral et al., 2024), and open defecation, which can expose individuals to mosquitoes (Agyemang-Badu et al., 2023; Gichuki et al., 2023; Olagunju, 2023). As with other vector-borne diseases, we included the use of mosquito nets and indoor spraying as potential prevention methods that could offer protective effects (Vargas Campos et al., 2025). At the individual level, we considered age and gender, which have been shown to be associated with mosquito exposure in several studies (Power et al., 2022).

**Table 1.** Socio-economic and demographic variables collected through household survey.

| Variables | Description |
| --- | --- |
| **Binary** |  |
| Rudimentary Housing | The house has both a rudimentary roof material (no roof, bamboo, thatch/palm leaves/leaves, clumps of grass, mats, and wooden planks) and rudimentary wall material (uncovered adobe, bamboo with mud, bamboo/cane/palm/trunk, and earth) |
| Secondary education or higher | The head of household has attended secondary school or higher |
| Cropland ownership | The household owns cropland |
| Livestock ownership | The household owns livestock |
| Household poultry ownership | The household owns poultry |
| Household Mosquito Net Ownership (all types) | The household reports having a mosquito net |
| Sufficient household Mosquito Net Ownership (LLIN) | The household owns one LLIN per two household members during the prior 24 months |
| Individual Mosquito Net Use (LLIN) | The individual slept under a LLIN during the prior 24 hours |
| Indoor residual spraying | The household received IRS in prior 12 months |
| Open defecation | The individual practices outside toilet habits |
| Outdoor occupation | The individual is over 14 years old and has one of the following outdoor occupations: agriculture and livestock farming, mining, day laborer |
| Unprotected water supply source | The household uses an unprotected water source |
| Male Sex | The individual is male |
| **Numeric** |  |
| Economic level | Wealth score of household based on asset ownership (ranging from 1 (lowest)-5 (highest)) |
| Household cropland area | Area of cropland owned by household (in hectares) |
| Total number of mammalian livestock | Quantity of mammalian livestock owned by family |
| Total number of poultry livestock | Quantity of poultry livestock owned by family |
| Age | Age of individual (in years) |

### Environmental factors

We collected environmental variables related to mosquito habitats, the characteristics of the mosquito life cycle, and the environment in which exposure takes place. These included climate variables, such as temperature, precipitation, and humidity, which influence the abundance and survival of *Culex* mosquitoes (Baril et al., 2023). We also collected data on potential mosquito larval habitats, including vegetation cover types, the amount of standing water in rice fields, the Topographic Wetness Index, and deforestation rates. Variables describing the population’s spatial structure, including population density, proximity to the National Park, proximity to public health centers, and proximity to paved and unpaved roads, were also included (García-Carrasco et al., 2023; Erazo et al., 2024; Gonçalves do Amaral et al., 2024; Vargas Campos et al., 2025). The full list of environmental variables used in this study is described in Table 2, along with details about their source and pre-processing methods.

**Table 2.** Description of environmental variables included in the initial variable collection.

| Variable | Methodology | Resolution | Time Period | Sources |
| --- | --- | --- | --- | --- |
| **Climatic** |  |  |  |  |
| Annual Minimum Land Surface Temperature (LST) (°C) | MODIS Land Surface Temperature, processed to replace missing data following Shiff et al. 2021 | Temporal/Spatial (1 km) | june 2020-june 2021 | (Shiff et al., 2021) |
| Precipitation of Wettest Month (bioclimatic) (mm) | None | Temporal/Spatial (~5 km) | june 2020-june 2021 | WordClim v2.1 (BIO13) |
| Annual Maximum Daily Relative Humidity (RH) (%) | None | Temporal/Spatial (~10km) | june 2020-june 2021 | ECMWF ERA5 |
| **Landuse** |  |  |  |  |
| Annual Maximum Percentage of Forest loss over 5 year (%) | None | Spatial (~30m) | 2017-2021 | (Hansen et al., 2013) |
| Average Normalized Difference Vegetation Index (NDVI) | Sentinel-2 Imagery processed via the Sen2Extract tool following George et al., 2024 | Temporal/Spatial (10m) | June 2020- June 2021 | (George et al., 2024) |
| Percentage of Forest around household (%) | Semi-supervised classification of 2019 Sentinel-2 imagery following Dezetraka (QGIS) | Spatial (10m) | 2019 | (Ihantamalala et al., 2020) |
| Percentage of Residential area around household (%) | Semi-supervised classification of 2019 Sentinel-2 imagery following Dezetraka (QGIS) | Spatial (10m) | 2019 | (Ihantamalala et al., 2020) |
| Percentage of Waterbodies around household (%) | Semi-supervised classification of 2019 Sentinel-2 imagery following Dezetraka (QGIS) | Spatial (10m) | 2019 | (Ihantamalala et al., 2020) |
| Percentage Ricefields area around household (%) | Semi-supervised classification of 2019 Sentinel-2 imagery following Dezetraka (QGIS) | Spatial (10m) | 2019 | (Ihantamalala et al., 2020) |
| Percentage of Savannah area around household (%) | Semi-supervised classification of 2019 Sentinel-2 imagery following Dezetraka (QGIS) | Spatial (10m) | 2019 | (Ihantamalala et al., 2020) |
| Average Maximum of flooded rice fields during primary agricultural season (%) | Sentinel-1 imagery processed following Randriamihaja et al. 2025 | Temporal/Spatial (10m) | june 2020- june 2021 | (Randriamihaja et al., 2025) |
| Average Maximum of flooded rice fields during secondary agricultural season (%) | Sentinel-1 imagery processed following Randriamihaja et al. 2025 | Temporal/Spatial (10m) | june 2020- june 2021 | (Randriamihaja et al., 2025) |
| **Topography** |  |  |  |  |
| Average elevation (m) | SRTM Digital Elevation Model | Spatial (30m) |  | NASA SRTM |
| Average Topographic Wetness Index (TWI) | Calculated from SRTM DEM following Evans et al. 2021 | Spatial (30m) | 2021 | (Evans et al., 2021) |
| **Infrastructure** |  |  |  |  |
| Population density (buildings/km2) | Mean number of buildings within 100m radius of each household | Spatial | 2019 | OpenStreetMap |
| Distance to nearest primary health centers (PHC) (km) | Calculated on OpenStreetMap routing data for each village via ORSM algorithm in R (package, orsm, v3.5.0) | Spatial | 2019 | OpenStreetMap |
| Distance to Ranomafana Park (km) | Calculated on OpenStreetMap routing data for each village via ORSM algorithm e in R (package, orsm, v3.5.0) | Spatial | 2019 | OpenStreetMap |
| Distance between household and the nearest major paved road (km) | Calculated on OpenStreetMap routing data for each village via ORSM algorithm in R (package, orsm, v3.5.0) | Spatial | 2019 | OpenStreetMap |
| Distance between household and the nearest unpaved road (km) | Calculated on OpenStreetMap routing data for each village via ORSM algorithm in R (package, orsm, v3.5.0) | Spatial | 2019 | OpenStreetMap |
| Distance between household and the nearest footpath (km) | Calculated on OpenStreetMap routing data for each village via ORSM algorithm in R (package, orsm, v3.5.0) | Spatial | 2019 | OpenStreetMap |

Variables at a monthly resolution (e.g., temperature, NDVI, rice field flooding), were aggregated to annualized summary statistics covering the period from June 2020 to June 2021 following Table 2. Variables derived from the WorldClim model were extracted from the 2010-2020 dataset and deforestation-related variables cover the five years preceding the survey (2017–2021). All other variables were constant. All spatial variables were extracted using a 500m buffer zone surrounding the villages included in the IHOPE survey (N = 477), then spatially averaged and assigned to all individuals residing in these villages, using an approach similar to that used by Giesen et al. (Giesen et al., 2023). All extractions and processing were performed using R (v. 4.5.0) and QGIS (v. 3.22.4).

## Data analysis

### Estimation of seropositive individuals

MFI values for both WNV E and NS1 proteins were log-transformed (Figure 2A). To account for experimental variability inherent to the Luminex assay, including operator and plate-related effects. 21 out of 67 plates were excluded when the coefficient of variation (CV = standard deviation/mean) of positive control duplicates exceeded 0.2 (see Appendix, S1 Fig), or when the mean positive control signal deviated more than two standard deviations from the overall distribution of positive controls (see Appendix, S1 Fig). A Gaussian mixture model (GMM) was then used to classify log-transformed MFI values into two groups representing individuals who were positive or negative for the antigen, using a threshold three Standard deviation (SD) above the mean of the presumed seronegative component to minimize false positives, following Cox et al. and Domingues et al. with R package *mixdist* (v. 0.5-5) (Figure 2A) (Domingues et al., 2021; Cox et al., 2022; Domingues et al., 2024). Cut-offs using a threshold two SD above the mean of the negative group yielded similar spatial patterns (see Appendix, Fig. S2). When applied to both antigens, this led to four total classifications: probable WNV (E+NS1+), false positive for WNV (E-NS1+), possible exposure to another orthoflavivirus (E+NS1-), and negative for any orthoflavivirus exposure (E-NS1-) (Figure 2C). We defined the probable WNV group (positive for both E and NS1) as seropositive for the rest of this study.

**Figure 2.**
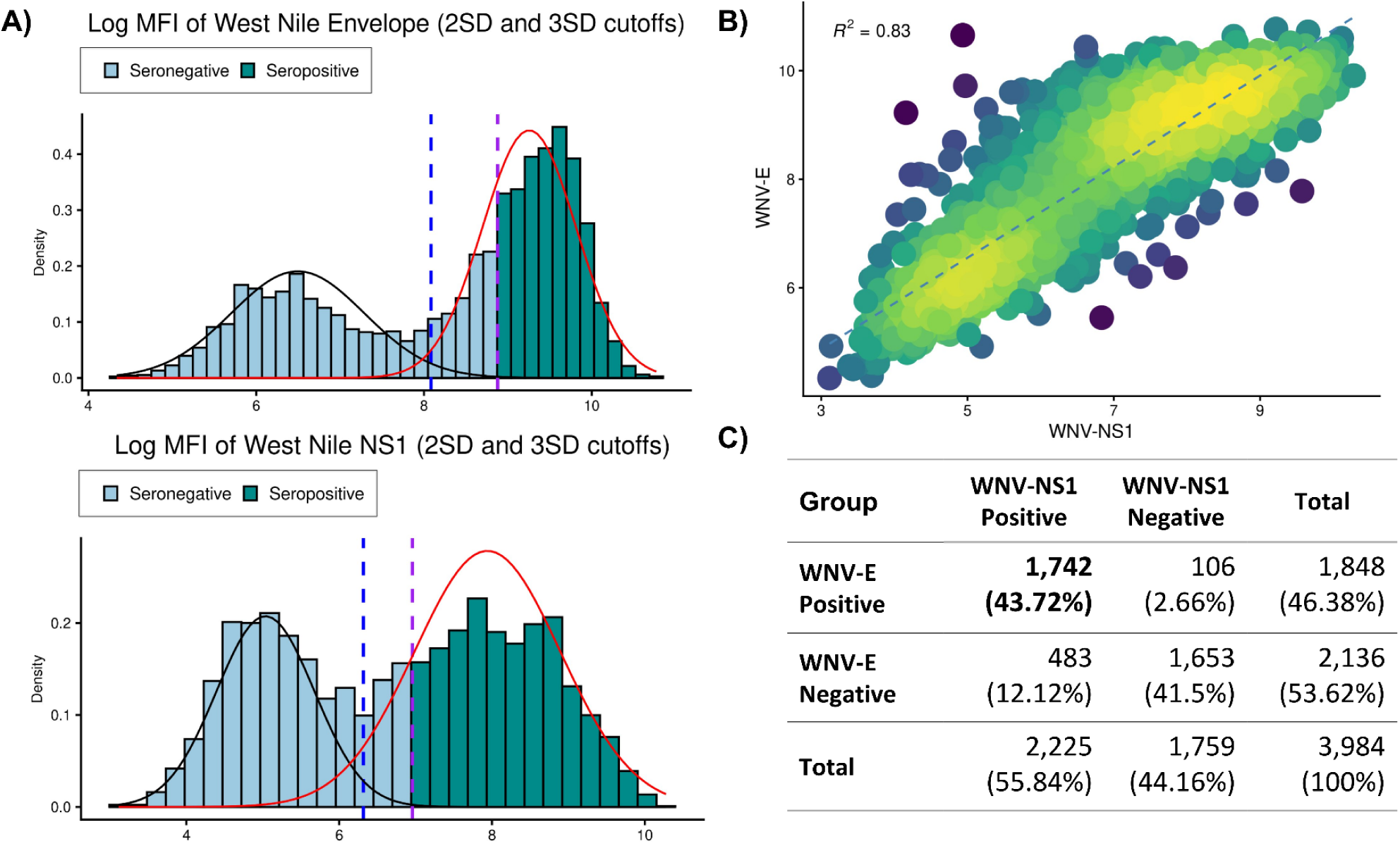
Distribution of antibody reactivity to WNV envelope protein (E) and nonstructural protein 1 (NS1). A) Distribution of two classes with a GMM model for each protein: positive and negative for WNV infection. Dotted vertical lines mark the cut-offs for seropositivity using 2 (blue) and 3 (purple) Standard deviation (SD) above the mean of the presumed negative group (black). The green bars represent seropositive individuals. B) Relationship between antibody binding intensity to WNV-E and WNV-NS1. C) Summary of seropositivity classifications for WNV-E and WNV-NS1.

### Regression analysis

We conducted statistical regression analyses to identify environmental, socioeconomic, and demographic factors associated with WNV exposure (see Appendix, S3 and S4 Fig). Given potential uncertainty in the cut-offs for WNV seropositivity, this included two modeling exercises: one considering the corrected MFI values of the WNV-E protein and another considering seropositivity data based on the GMM results defined above (Figure 2). We applied the same modeling approach for both exercises. First, we inspected the initial dataset of 38 explanatory variables to determine collinearity between them and removed those variables that were highly collinear (Spearman ***ρ*** > 0.7) (see Appendix, S3 Fig). Three environmental variables were removed (average elevation, distance to Ranomafana Park, and household poultry ownership), while outdoor occupation, which was collinear with age, was kept, leaving 35 variables (see Appendix, S4 Fig). We constructed hierarchical models, taking into account the design of the survey data with random intercepts for the villages nested within each cluster. For the seropositivity analysis, we used a generalized linear mixed model with a binomial family and log link. For analysis of MFI values, we used a linear mixed model with log-transformed MFI values as the outcome variable. To aid in model convergence and interpretation, we standardized all explanatory variables by subtracting the mean and dividing by the standard deviation. We created the final model using a backward selection procedure based on the Akaike information criterion (AIC) value (Sutherland et al., 2007), starting with a complete set of 35 variables and fixing the random effects. We performed statistical regression analyses using the *glmmTMB* package, v. 1.1.11 (Brooks et al., 2025), variable selection using the *buildmer* package, v. 2.12 (Voeten, 2019). All analyses were performed using R statistical software (v. 4.5.0).

## Ethics statement

French and Malagasy questionnaires used in the cohort, as well as data collection protocols, were standardized and validated for Madagascar during previous national surveys carried out by INSTAT. The study was approved by the Madagascar National Ethics Committee and Harvard Medical School IRB, including amendments for changes in the 2021 surveys. Before data collection, written informed consent was obtained from the household representative after explaining the study procedures, including questionnaires and biological sampling of household members. Individuals aged ≥15 years provided informed consent for individual interviews and separate written informed consent for biological sample collection. For children aged <15 years, the household representative additionally provided specific written consent for biological sampling of each child, while children aged 7–14 years provided separate written assent. For adolescents aged 15–17 years, written consent for biological sampling was obtained directly from the participants, following the household-level consent process, without a separate written parental consent form.

## Results

### West Nile virus seroprevalence in Ifanadiana District

A total of 3,984 samples were analyzed, after removing samples due to high variability in Luminex assay quality. The MFI values for both WNV-E and WNV-NS1 antigens showed a bimodal, Gaussian mixture distribution, with values ranging from 6.82 to 9.45 for the E protein and from 5.54 to 8.10 for the NS1 protein (Figure 2A). Comparison between MFI values of WNV-E and WNV-NS1 revealed a strong correlation (R^2= 0.83) (Figure 2B). 46.38% of samples were seropositive for E, 55.84% for NS1 (see Methods), and 43.72% were positive for both antigens, which were considered seropositive for WNV (Figure 2C). Seroprevalence increased rapidly with age, especially in clusters with medium or high seroprevalence (see Appendix, S5 Fig).

### Spatial modeling to identify factors associated with West Nile virus exposure

We examined the spatial distribution of WNV seroprevalence for the E and NS1 proteins, as well as for both proteins combined (WNV positive) (Figure 3). Seropositivity to both the E and NS1 proteins combined showed a clear and consistent spatial structure, with higher but more heterogeneous patterns of seroprevalence in the northern and southern parts of the district, and lower seroprevalence in the center, with seroprevalence ranging from about 9% to nearly 100% at the cluster-level (Figure 3A). Similarly, WNV seroprevalence for each protein individually (E and NS1) had a pronounced spatial structure, with cluster-level seroprevalence ranging from 9% to 100% (Figure 3B-C).

**Figure 3.**
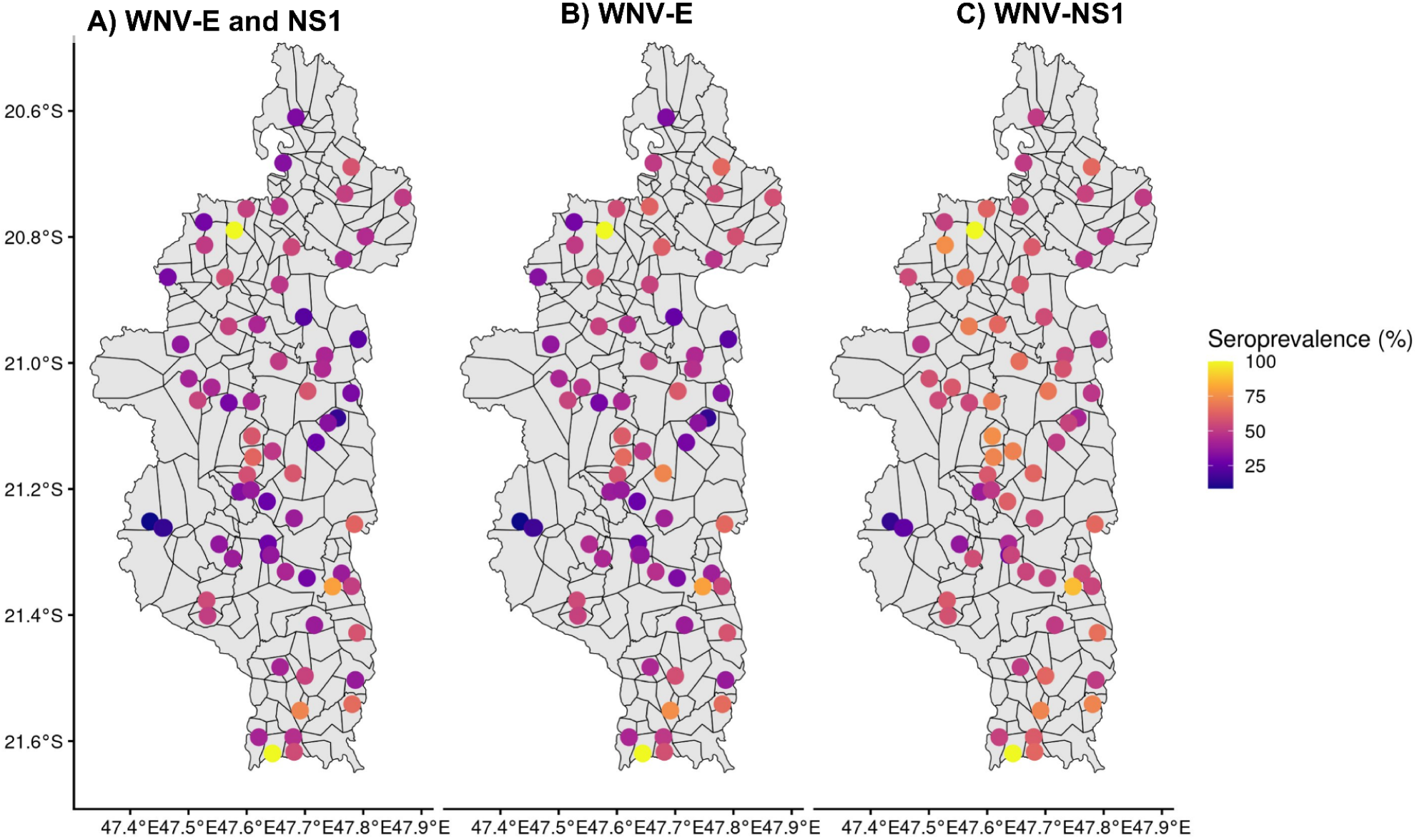
Spatial distribution of the proportion of seropositivity by WNV antigen. A) WNV-E and NS1 combined, B) WNV-E, and C) WNV-NS1. A similar figure showing the MFI value measured on the WNV-E and NS1 protein is available in the supplementary material (see Appendix, S6 Fig). Sources: HumData Madagascar (https://data.humdata.org/dataset/cod-ab-mdg) used for administrative boundaries.

The statistical model retained eight environmental variables and three socio-economic and demographic variables associated with WNV seroprevalence (Figure 4). WNV seropositivity was positively associated with both rainfall (OR=1.31, 95% CI: 1.13-1.52) and NDVI (OR=1.14, 95% CI: 1.02-1.28). The proportion of forested and savannah land cover (OR=0.73, 95% CI: 0.61-0.87 and OR=0.83, 95% CI: 0.67-1.03), measured within buffers surrounding households, were negatively associated with seropositivity. Seropositivity was also negatively associated with household proximity to primary health centers (PHCs) (OR=0.89, 95% CI: 0.79-0.99). Conversely, seropositivity tended to be higher in more remote areas located far from the paved national road (OR=1.25, 95% CI: 1.08-1.45), and in areas with fewer residential structures surrounding households (i.e., located in less densely populated areas). Similarly, households that owned more agricultural land (OR=1.08, 95% CI: 1.00-1.16) and individuals with outdoor activities (OR=2.00, 95% CI: 1.68-2.39) as their primary occupation (agriculture, etc.) had a higher risk of seropositivity. Mosquito net use had a protective effect (OR=0.77, 95% CI: 0.63-0.94), while WNV seropositivity increased with increasing age of the individual (OR=1.30, 95% CI: 1.19-1.42) (see Appendix, S5 Fig).

**Figure 4.**
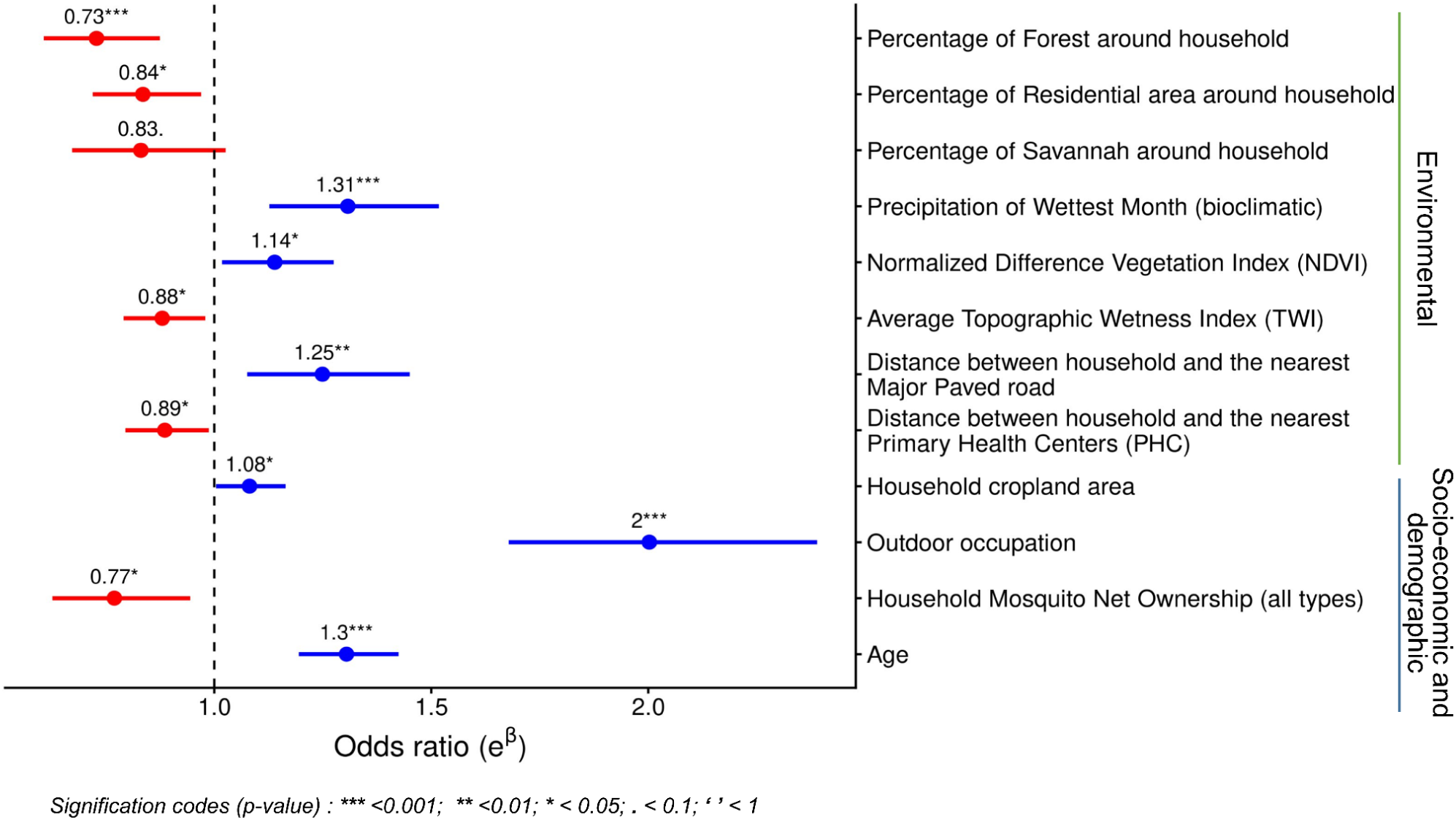
Odds ratios (error bars represent 95% CI) of the environmental, socioeconomic, and demographic variables associated with WNV seropositivity in Ifanadiana District, Madagascar, 2021. A similar figure is available for the supplemental analysis on WNV-E MFI values using the GLMM model with a Gaussian family and random effects, as described in the Methodology section (see Appendix, S7 Fig).

## Discussion

West Nile virus (WNV) is a febrile-causing arbovirus with a global presence that has recently been recognized as a neglected disease in tropical regions, outside its initial geographic zone of concern in the Northern Hemisphere (Ronca et al., 2021). Prevalence rates are particularly high in sub-Saharan Africa (SSA), where the environmental, socioeconomic, and demographic conditions are suitable for arbovirus transmission (Sule et al., 2018; Klingelhöfer et al., 2023; García-Carrasco et al., 2023; Mencattelli et al., 2023; Giesen et al., 2023; Vargas Campos et al., 2025). Identifying the factors associated with the distribution of the virus in tropical rural areas is essential to understanding its spread, reducing the risk of transmission and mitigating its impact on human health. In this study, we estimated seroprevalence rates of WNV exposure in humans in a rural district of southeastern Madagascar, and identified the socio-ecological factors influencing the local distribution of the disease. We observed a high prevalence of antibodies against WNV antigens, increasing with age, suggesting that WNV transmission is endemic in the study area. Seroprevalence rates showed a pronounced spatial pattern, with the highest rates observed in less densely populated rural areas located farther from the primary transportation route. Local heterogeneity in environmental and climatic factors related to mosquito proliferation (e.g. forest cover, NDVI, rainfall) was associated with spatial patterns in the risk of exposure, while socio-demographic characteristics (e.g. outdoor occupations, bednet use, age) modulated individuals’ risk. These results highlight the value of fine-scale seroepidemiology analyses to obtain actionable insights into high-risk areas and key factors that can help guide prevention and control of WNV transmission in this and similar low-resource settings.

Our results suggest that WNV circulates in Ifanadiana district, with the more rural northern and southern areas of the district experiencing the highest seroprevalence rates (Figure 3). More than 40% of individuals tested (N=3,984) showed serological evidence of previous WNV infection. This seroprevalence rate is slightly higher than those found in other sub-Saharan African countries and in other studies conducted in Madagascar. Using similar serological methods, based on a multiplex ELISA targeting IgG antibodies, seroprevalence was estimated at 39.1% in Mali between 2009 and 2013 (Mencattelli et al., 2022), and 27.2% in Gabon (Sule et al., 2018). In Madagascar, studies have previously reported seroprevalences of 27.1% (between 24.4% and 29.8%) for WNV-NS1 in five provinces throughout the country (Rakotomalala et al., 2023), and 25% (between 17.9% and 32.1%) in the southeast (Broban et al., 2023). In comparison, our study demonstrates a wide range of cluster-level seroprevalence rates, including some clusters with seroprevalence near 100%, reflecting substantial spatial heterogeneity across a single district. These findings underscore the added value of such high-resolution (sub-district) surveys, in addition to surveys conducted at the national level, in order to identify local WNV hotspots and at-risk groups, which can inform surveillance and control measures to limit the spread of the virus within local communities.

In our setting, proximity to the national road was associated with reduced risk of WNV exposure, and communities in more remote areas had higher exposure to the virus. WNV is transmitted by *Culex* mosquitoes, which are often found in peri-urban areas (Bravo-Barriga et al., 2017), but can be abundant in more remote rural areas. Environmental conditions positively associated with WNV exposure here, such as heavy rainfall, or NDVI, can amplify the spread and growth of many mosquito species, including *Culex* (Lockaby et al., 2016; Ateutchia Ngouanet et al., 2022; Wang et al., 2024; Gonçalves do Amaral et al., 2024; Vargas Campos et al., 2025). Conversely, the protective effect seen for household near residential areas and those with higher savannah and forest cover, which is consistent with previous studies (Lockaby et al., 2016; Sule et al., 2018; Giesen et al., 2023), may reflect complex interactions between habitat characteristics, vector ecology, and host availability. Residential environments may support mosquito populations with limited WNV amplification potential if infection prevalence among adult mosquitoes is low or if contact with competent reservoir hosts is reduced. Similarly, forest and savannah ecosystems may provide cooler, more humid, and shaded microhabitats that influence mosquito survival, resting behavior, and host-seeking activity (Xia et al., 2021; Coulibaly et al., 2025), potentially maintaining mosquito-avian transmission cycles while limiting opportunities for spillover to humans and livestock. When *Culex* mosquitoes remain primarily associated with avian reservoir hosts, WNV circulation may be sustained within the enzootic cycle, thereby reducing opportunities for spillover to mammalian hosts (Kilpatrick et al., 2006; Hamer et al., 2009). Surprisingly, after accounting for these environmental, topographic and climatic factors, the average topographic wetness index (TWI), reflecting how water accumulates in areas with elevation differences, had a protective effect against WNV exposure in our setting, differing from findings in other tropical countries (Ukawuba & Shaman, 2018; Taheri et al., 2025). In Ifanadiana, TWI was highest in low-lying rice fields, whose frequent flooding may result in unsuitable larval habitats for *Cx. quinquefasciatus* mosquitoes (Kovach & Kilpatrick, 2018). In addition to hyper-local variation in the physical environment, we also considered the local context with regards to geographical isolation by including multiple measures relevant in the district. The combination of geographical isolation and mosquito-favourable environmental conditions (e.g., rainfall and NDVI) can increase WNV exposure for whole communities, while behaviours and daily activities, especially intensive agriculture, may further modify individuals’ risk of exposure.

In addition to community-level risk factors, individual-level characteristics and behaviours further modulated WNV exposure risk in our setting. Several occupations common in rural areas of Madagascar and elsewhere, such as agriculture, livestock farming, mining, and outdoor day labor, were associated with increased WNV exposure, probably due to the expansion of cultivated land, which has been shown to create habitats favorable to vectors and contribute to WNV transmission (Lu et al., 2024; Odigie et al., 2024; Vargas Campos et al., 2025). The use of insecticide-treated mosquito nets (LLINs) was identified as a protective measure against WNV, in agreement with findings from other settings (Vargas Campos et al., 2025). Although primarily used for malaria prevention, whose vectors bite during the night, LLINs may also protect against crepuscular biters such as Culex mosquitoes through their insecticidal effects and if the human population sleeps under protective nets as early as dusk, as is the case in many rural areas of Madagascar. Finally, the odds of being seropositive increased with age (see Appendix, S5 Fig), in line with what has been previously observed (Kalantari et al., 2019; Olufemi et al., 2021; Rakotomalala et al., 2023), consistent with increased cumulative exposure in an endemic setting.

This study has several limitations. Collection of blood for serological analyses was done on dried blood spot (DBS) samples, which introduces potential variation during spotting, transport, storage, and elution steps. Although this might have introduced some heterogeneity in MFI results, this sampling method was necessary given the remote rural areas where the survey was conducted. Panels of confirmed true positive or negative cases of WNV were not available to assess the performance of the GMM classification model but clear bimodal distribution of MFI values and strongly correlated responses to two distinct WNV antigens provides confidence that the GMM appropriately classified responses. Moreover, the classification with two SD cutoffs resulted in a similar spatial seroprevalence pattern, also suggesting that the classification was robust (see Appendix, S2 Fig). While cross-reactivity with other orthoflaviviruses could contribute to some misclassification, the results of this study demonstrate a similar estimated seroprevalence beyond the studied district of Madagascar (Broban et al., 2023; Rakotomalala et al., 2023), suggesting the seroprevalence estimate is realistic for the region. Environmental data was extracted within a radius of 500 m of each village, assuming that most *Culex* mosquitoes are found near domestic sites (Bravo-Barriga et al., 2017). However, no entomological survey has been conducted in the district (although see M. L. Tantely et al 2016, 2017, 2024 for studies done elsewhere in the country) to identify the specific species of *Culex* present or their WNV positivity rate (M. L. Tantely et al., 2016; L. M. Tantely et al., 2017; M. L. Tantely et al., 2024), so exposure to mosquitoes could occur further from residential areas than was assumed. Finally, the lack of longitudinal follow-up does not allow us to directly measure changes in seropositivity over time or accurately estimate the time of exposure to WNV. However, seropositivity rates of 15,8% in children under-five and increased seropositivity in older age groups suggest both historic and recent circulation of WNV in the District.

In conclusion, this study found that environmental and climatic determinants of WNV exposure, particularly humid, vegetated landcover, are modulated by individuals’ occupations and preventative behaviors to create heterogeneous spatial risk patterns on a fine-scale. These results enable the accurate identification of areas and population groups vulnerable to WNV exposure. Integrating other arboviruses transmitted by different vectors (e.g. DENV, Chikungunya virus) into future analyses would provide a deeper understanding of the ecology of local viral co-circulation dynamics, refine the identification of infection-associated factors, and strengthen prevention and control strategies adapted to the local context.

## Data availability

Data are available upon request to the address. Analysis code is available on github (https://github.com/mauricianot/wnv.git).

## Supplementary data

**Supporting Information Appendix.** It contains 7 supplementary figures (Figures S1-S7).

## Author contributions

**Conceptualization:** Mauricianot Randriamihaja, Michelle V Evans, Andres Garchitorena

**Data curation:** Ezra Mitsinjoniala, Hobisoa Razanadranaivo, Benedicte Razafinjato, Ann C. Miller

**Formal analysis:** Lova Tsikiniaina Rasoloharimanana, Matthieu Schoenhals, Stephen J. Popper, Sandra Bos, Sruthi Veeragandham, Pierre Garneret, Felana Angella Ihantamalala, Mauricianot Randriamihaja, Michelle V Evans

**Funding acquisition:** Andres Garchitorena, Veronique Chevalier

**Investigation:** Lova Tsikiniaina Rasoloharimanana, Matthieu Schoenhals, Rado JL Rakotonanahary, Stephen J. Popper, Karen E. Finnegan, Mauricianot Randriamihaja, Matthew H. Bonds

**Methodology:** Stephen J. Popper, Pierre Garneret, Mauricianot Randriamihaja, Michelle V Evans, Andres Garchitorena

**Project administration:** Andres Garchitorena, Veronique Chevalier

**Software:** Sruthi Veeragandham, Pierre Garneret, Mauricianot Randriamihaja

**Supervision:** Matthieu Schoenhals, Stephen J. Popper, Michelle V Evans, Andres Garchitorena

**Validation:** Mauricianot Randriamihaja, Lova Tsikiniaina Rasoloharimanana, Stephen J. Popper, Michelle V Evans

**Visualization:** Stephen J. Popper, Pierre Garneret, Mauricianot Randriamihaja

**Writing – original draft preparation:** Mauricianot Randriamihaja, Michelle V Evans, Andres Garchitorena

**Writing – review & editing:** Mauricianot Randriamihaja, Lova Tsikiniaina Rasoloharimanana, Sruthi Veeragandham, Ann C. Miller, Karen E. Finnegan, Sandra Bos, Benedicte Razafinjato, Ezra Mitsinjoniala, Jacques Aubin Kotchofa, Pierre Garneret, Hobisoa Razanadranaivo, Felana Angella Ihantamalala, Rado JL Rakotonanahary, Veronique Chevalier, Matthew H. Bonds, Matthieu Schoenhals, Michelle V Evans, Stephen J. Popper and Andres Garchitorena

## Acknowledgments

We would like to express our sincere gratitude to the NGO Pivot’s Medical teams for their support during the 2021 wave of the IHOPE longitudinal cohort study. We would also like to thank the field teams at INSTAT for their effort and quality work during the survey, and to all the survey participants for their contribution through valuable time and samples.

