## Supporting Information Appendix for "Spatial modeling of risk factors for human exposure to West Nile virus in rural Madagascar"

**S1 Fig. A) Coefficient of variation (CV) of duplicate positive controls for WNV antigens (envelope and NS1) across Luminex plates** (plates with CV > 0.2 are labelled). **B) Standardized mean log (MFI) values of positive controls for WNV antigens (envelope and NS1) across plates** (plates with values deviating by more than 2 Standard deviations from the overall mean are labelled).

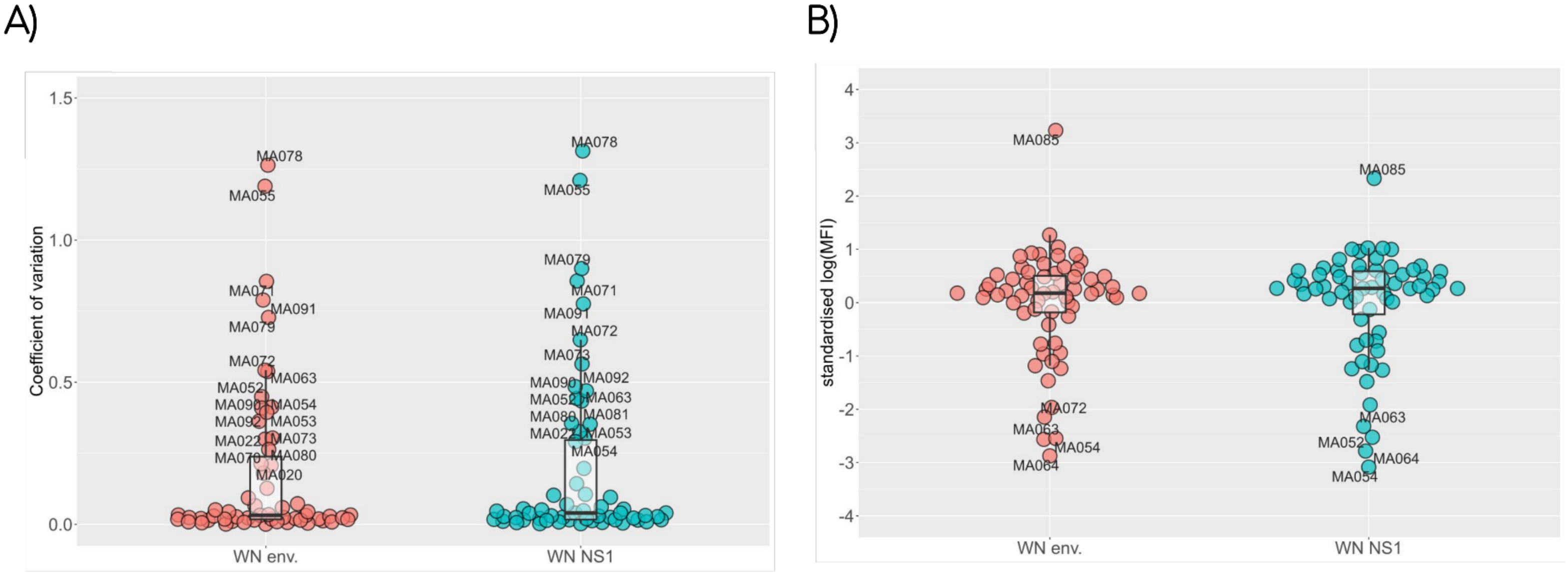

**S2 Fig. A comparison of the spatial distribution of WNV seroprevalence using two and three SD cut-offs.**

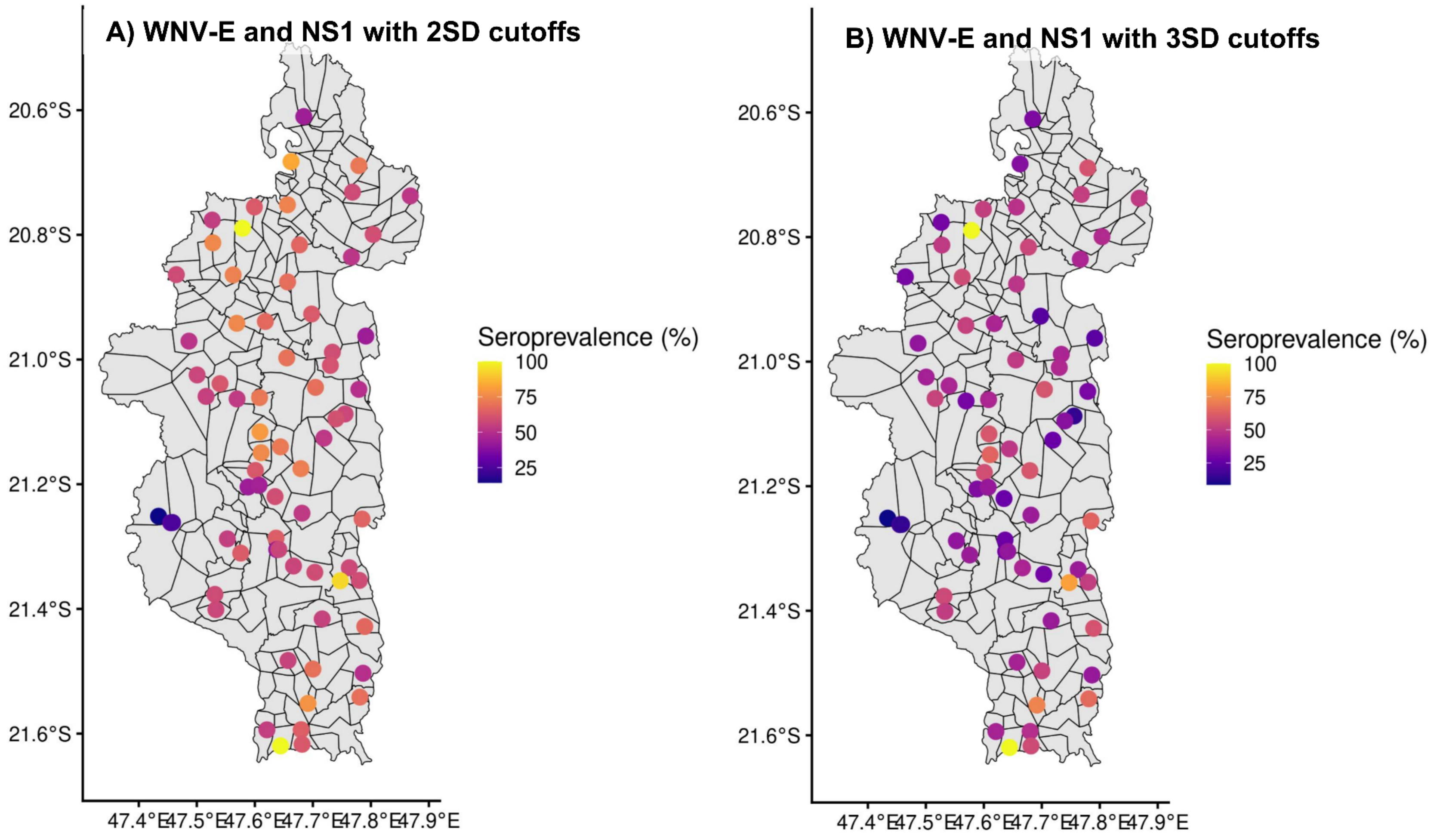

**S3 Fig. Correlation matrix of the 38 independent variables (environmental, socioeconomic, and demographic) used to identify and remove variables with a correlation coefficient of at least 0.7.**

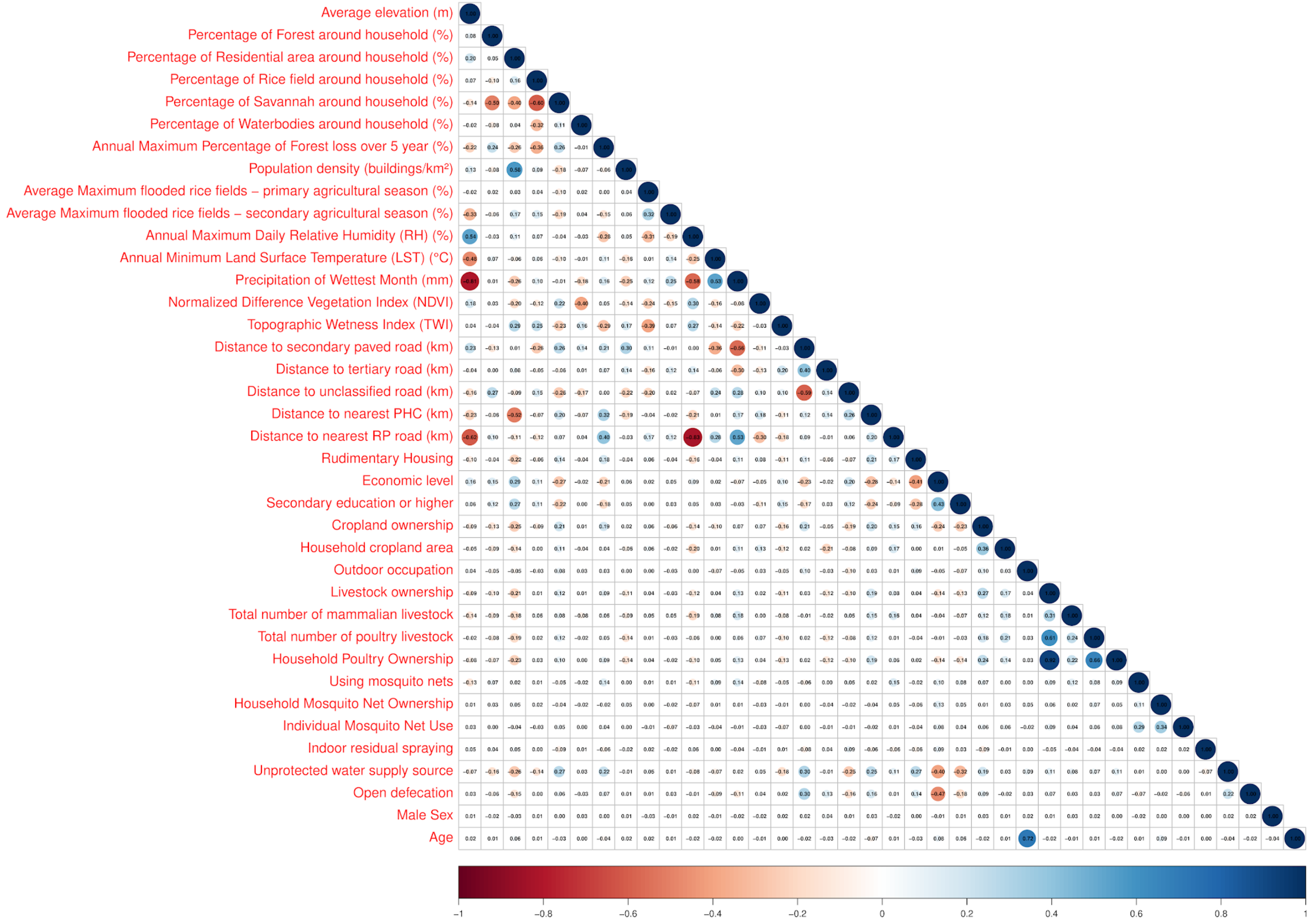

**S4 Fig. Correlation matrix of the 35 independent variables (environmental, socioeconomic, and demographic) selected with a correlation coefficient of less than 0.7, except between the variables Age and High exposure occupation (42.50% of the individuals under the age of 14 years are considered students).**

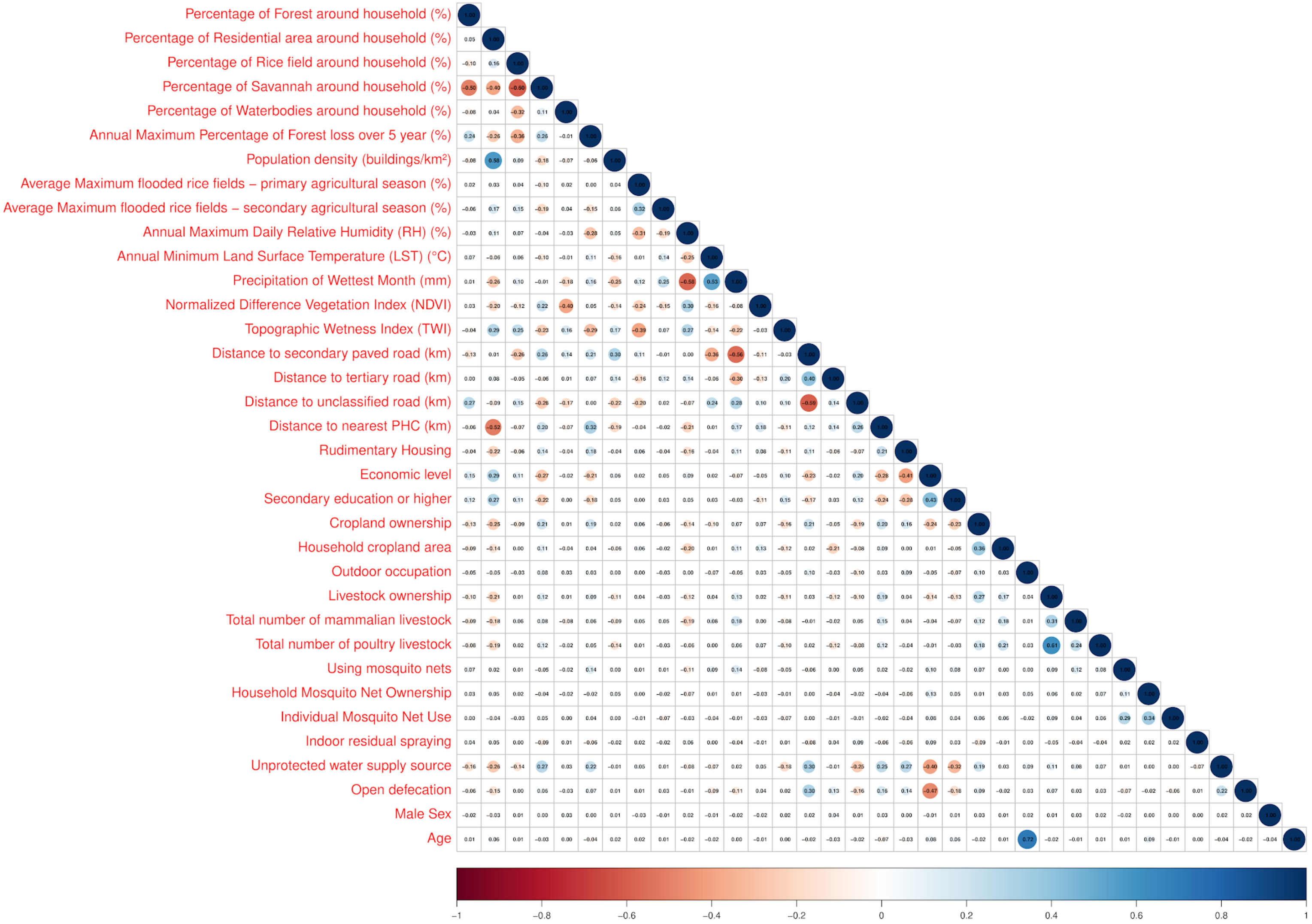

**S5 Fig. Distribution of WNV seroprevalence by age category and cluster-level risk (low, medium, and high).** A) Age-specific seroprevalence for the entire district. B) Age-specific seroprevalence curves for high, medium, and low prevalence clusters. Each group shows an increase in exposure with age (infant, child, young, teen/young adult, adult, and older adult).

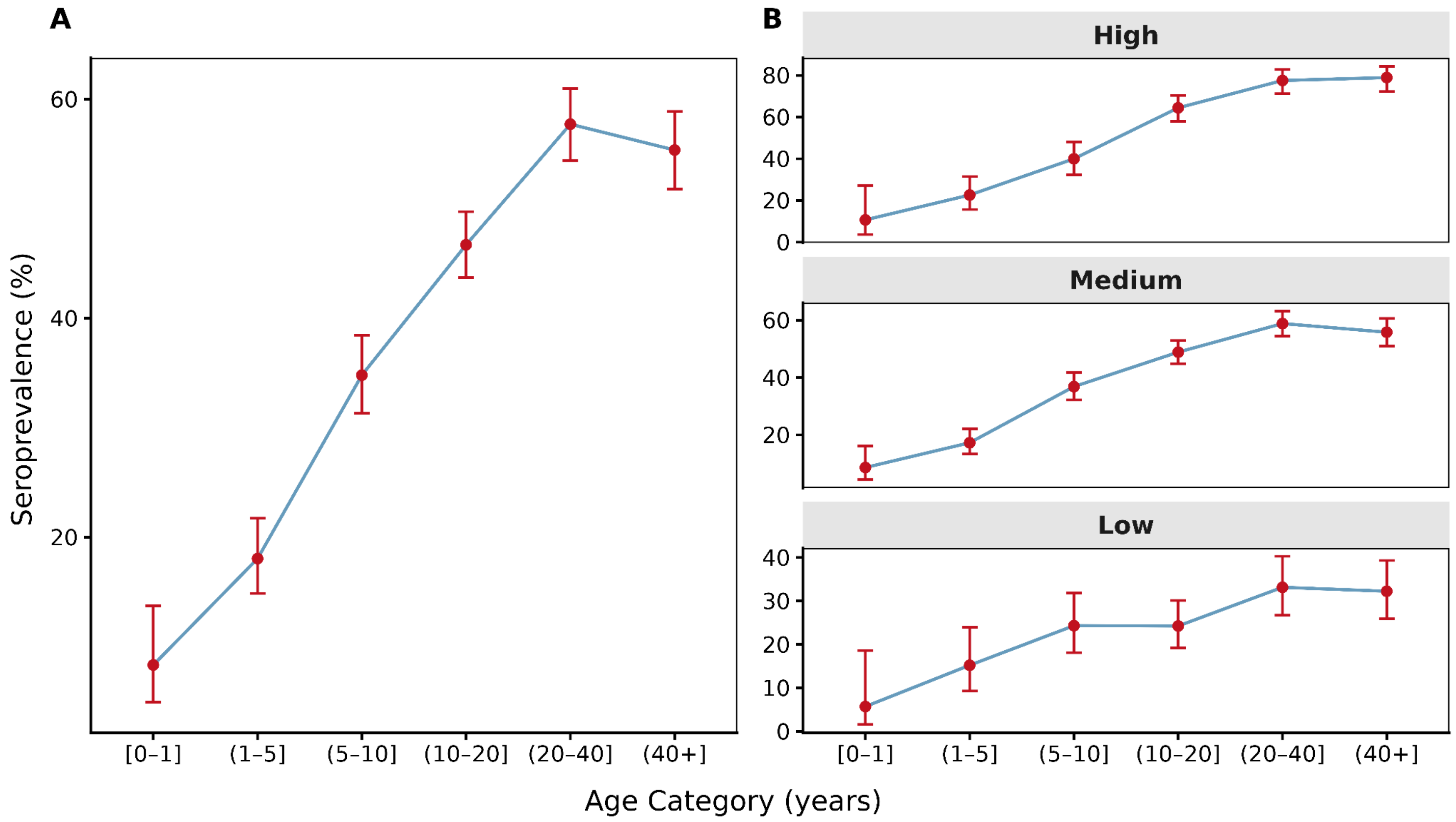

**S6 Fig. Spatial distribution of the proportion of mean fluorescence intensity (MFI) of the West Nile virus (WNV) to the envelope protein (E), and nonstructural protein 1 (NS1).** A) WNV envelope protein (WNV-E), and B) WNV nonstructural protein 1 (WNV-NS1).

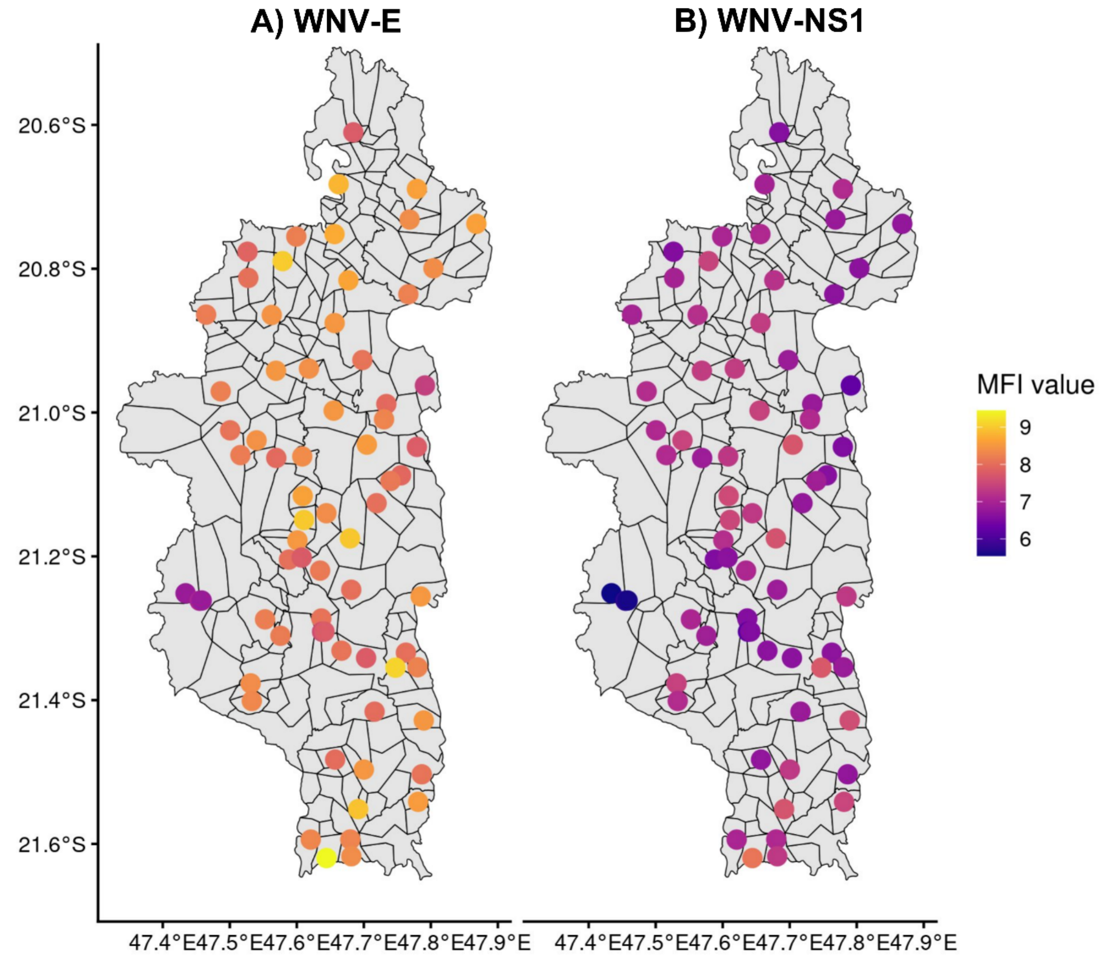

**S7 Fig. Odds ratios (error bars represent 95% CI) of the environmental, socioeconomic, and demographic variables associated with West Nile virus (WNV) envelope protein mean fluorescence intensity (MFI) in Ifanadiana District.**

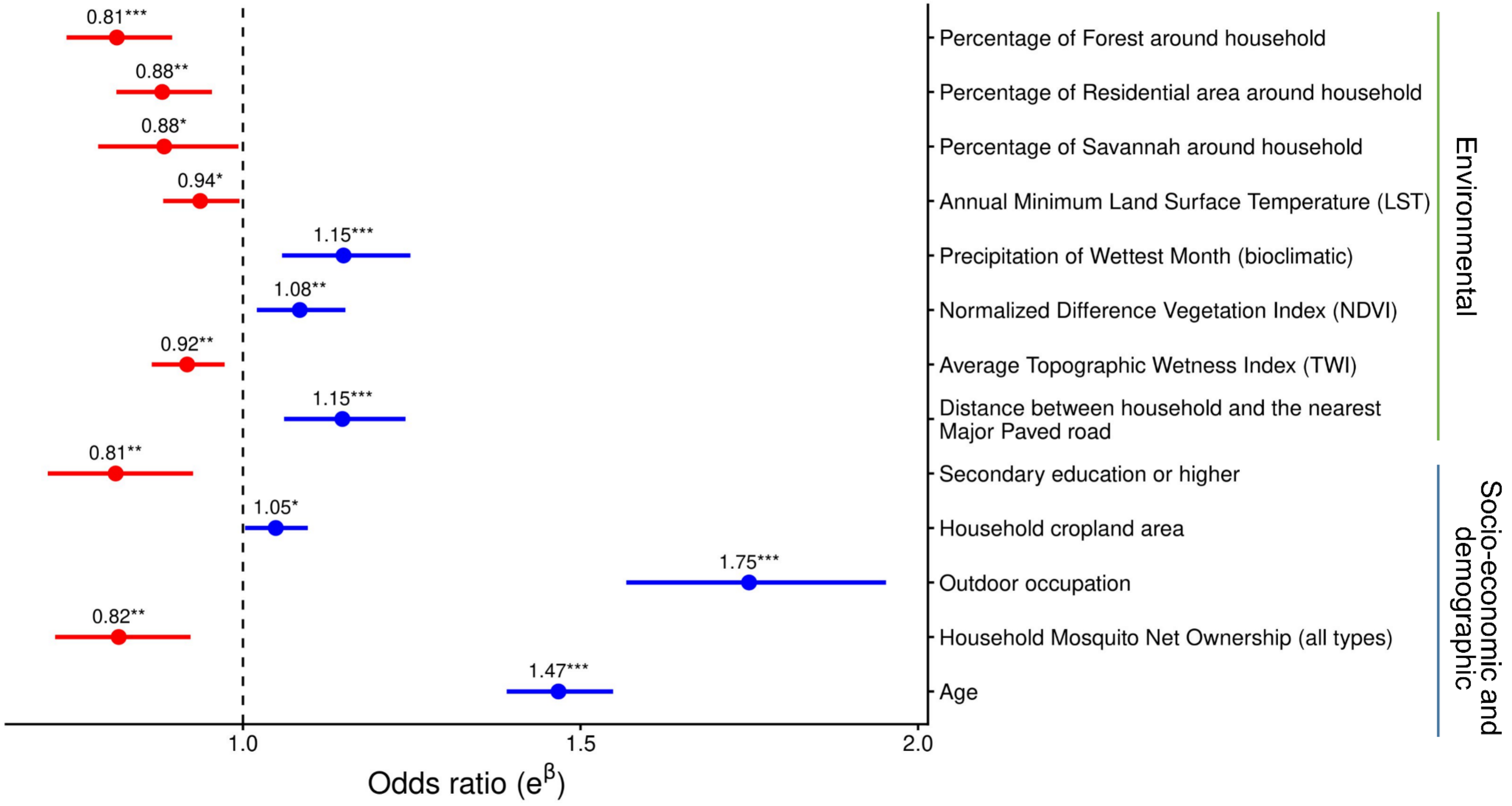


*Signification codes (p-value) : *** <0.001; ** <0.01; * < 0.05; . < 0.1; ‘ ’ < 1*
